# Performance of Heart Failure Clinical Prediction Models: A Systematic External Validation Study

**DOI:** 10.1101/2021.01.31.21250875

**Authors:** Jenica N. Upshaw, Jason Nelson, Benjamin Koethe, Jinny G. Park, Hannah McGinnes, Benjamin S. Wessler, Marvin A. Konstam, James E. Udelson, Ben Van Calster, David van Klaveren, Ewout Steyerberg, David M. Kent

## Abstract

**Background:** Most heart failure (HF) clinical prediction models (CPMs) have not been externally validated.

**Methods:** We performed a systematic review to identify CPMs predicting outcomes in HF, stratified by acute and chronic HF CPMs. External validations were performed using individual patient data from 8 large HF trials (1 acute, 7 chronic). CPM discrimination (c-statistic, % relative change in c-statistic), calibration (calibration slope, Harrell’s E, E90), and net benefit were evaluated for each CPM with and without recalibration.

**Results:** Of 135 HF CPMs screened, 24 (18%) were compatible with the population, predictors and outcomes to the trials and 42 external validations were performed (14 acute HF, 28 chronic HF). The median derivation c-statistic of acute HF CPMs was 0.76 (IQR, 0.75, 0.8), validation c-statistic was 0.67 (0.65, 0.68) and model-based c-statistic was 0.68 (0.66, 0.76), Hence, most of the apparent decrement in model performance was due to narrower case-mix in the validation cohort compared with the development cohort. The median derivation c-statistic for chronic HF CPMs was 0.76 (0.74, 0.8), validation c-statistic 0.61 (0.6, 0.63) and model-based c-statistic 0.68 (0.62, 0.71), suggesting that the decrement in model performance was only partially due to case-mix heterogeneity. Calibration was generally poor - median E (standardized by outcome rate) was 0.5 (0.4, 2.2) for acute HF CPMs and 0.5 (0.3, 0.7) for chronic HF CPMs. Updating the intercept alone led to a significant improvement in calibration in acute HF CPMs, but not in chronic HF CPMs. Net benefit analysis showed potential for harm in using CPMs when the decision threshold was not near the overall outcome rate but this improved with model recalibration.

**Conclusions:** Only a small minority of published CPMs contained variables and outcomes that were compatible with the clinical trial datasets. For acute HF CPMs, discrimination is largely preserved after adjusting for case-mix; however, the risk of net harm is substantial without model recalibration for both acute and chronic HF CPMs.

## Introduction

There are more than 6 million people living with heart failure (HF) in the United States.^1^ Individuals with HF experience significant morbidity due to symptoms, reduced functional capacity, frequent hospitalizations and are at increased risk for early mortality.^2-4^ While overall median survival is only 3-5 years, there is significant variation in risk of short-term morbidity and mortality.^5, 6^ Multiple patient characteristics including age, comorbidities such as atrial fibrillation and diabetes, measures of symptom severity, echocardiographic variables and laboratory values such as serum creatinine and sodium have been consistently associated with increased risk for mortality in individuals with HF.

Individualized outcome predictions may be useful to patients and providers to aid in: 1) discussions around prognosis, 2) decision making around numerous HF therapies such as transplant or left ventricular assist device, implantable cardioverter-defibrillators, HF disease management or palliative care or 3) research efforts including risk adjustment in observational studies, eligibility criteria for trials, or use to examine heterogeneous effects. Clinical prediction is improved by combining multiple prognostic variables in clinical prediction models (CPMs), which can provide patient-specific risk estimates. Multiple CPMs have been created for HF outcomes, most commonly the outcomes of all-cause mortality and/or hospitalization, and the use of CPMs for clinical decision making is endorsed by HF guidelines.^7, 8^

In prior research using the Tufts Predictive Analytics and Comparative Effectiveness (PACE) CPM Registry, a systematic collection of published cardiovascular CPMs, we reported that there has been a steady increase in new cardiovascular CPMs; however, most published CPMs have not been externally validated – i.e. performance tested in a separate cohort of patients distinct from the cohort in which the model was developed.^9^

While several HF CPMs have been externally validated in multiple cohorts, few CPMs have been independently externally validated – i.e. external validation performed by researchers unaffiliated with the initial model development. In addition, few studies have compared the performance of multiple CPMs in the same external validation cohort or applied a systematic approach to HF CPM external validation. Few external validations consistently report measures of discrimination, calibration and net benefit or explore the effect of recalibration on model performance. We aimed to review published HF CPMs and evaluate performance with and without CPM recalibration.

## Methods

Systematic Review of CPMs: This study used the Tufts PACE CPM Registry to identify HF CPMs. The registry has been previously reported and represents a systematic review of cardiovascular CPMs.^9^ We performed a PubMed search for English-language articles containing newly developed CPMs for cardiovascular disease from January 1990 through 2015 (search terms available in Supplement). This search was supplemented with reference lists of published articles. For this study, we restricted the population to CPMs reporting clinical outcomes, such as mortality or hospitalization, in individuals with established HF. CPMs were defined as models that provide a method to calculate or categorize an individual patients’ absolute risk for a binary outcome and includes at least 2 predictors. We did not restrict by outcome. For each CPM the following was extracted: derivation cohort demographic and clinical information (e.g. age, comorbidities, laboratory and imaging values), type of cohort (registry, clinical trial), cohort enrollment location and years, variable selection method (if described), variables in final model, CPM outcome definition(s), outcome event rate, events per included variable and whether the full risk equation was reported versus a risk score alone. Blinded double extraction was done on a random 10% sample of articles as a quality check. The registry was also queried for published external validations for the matched CPMs.

Validation Datasets: Publicly available clinical trial datasets^10-15^ were accessed using through the National Heart, Lung and Blood Institute via the Biologic Specimen and Data Repository Information Coordinating Center (BioLINCC) and supplemented with two industry-sponsored clinical trials^16, 17^ (Table 1). The clinical trial cohorts were restricted to HF with reduced left ventricular ejection fraction (LVEF) in 7 trials and HF with preserved LVEF in one cohort^10^. One trial included individuals with acute decompensation of chronic HF^17^ while the other 7 trials enrolled outpatients with chronic HF. Cohort enrollment years ranged from 1986 to 2012 (median trial enrollment year was 2005).

**Table 1:**
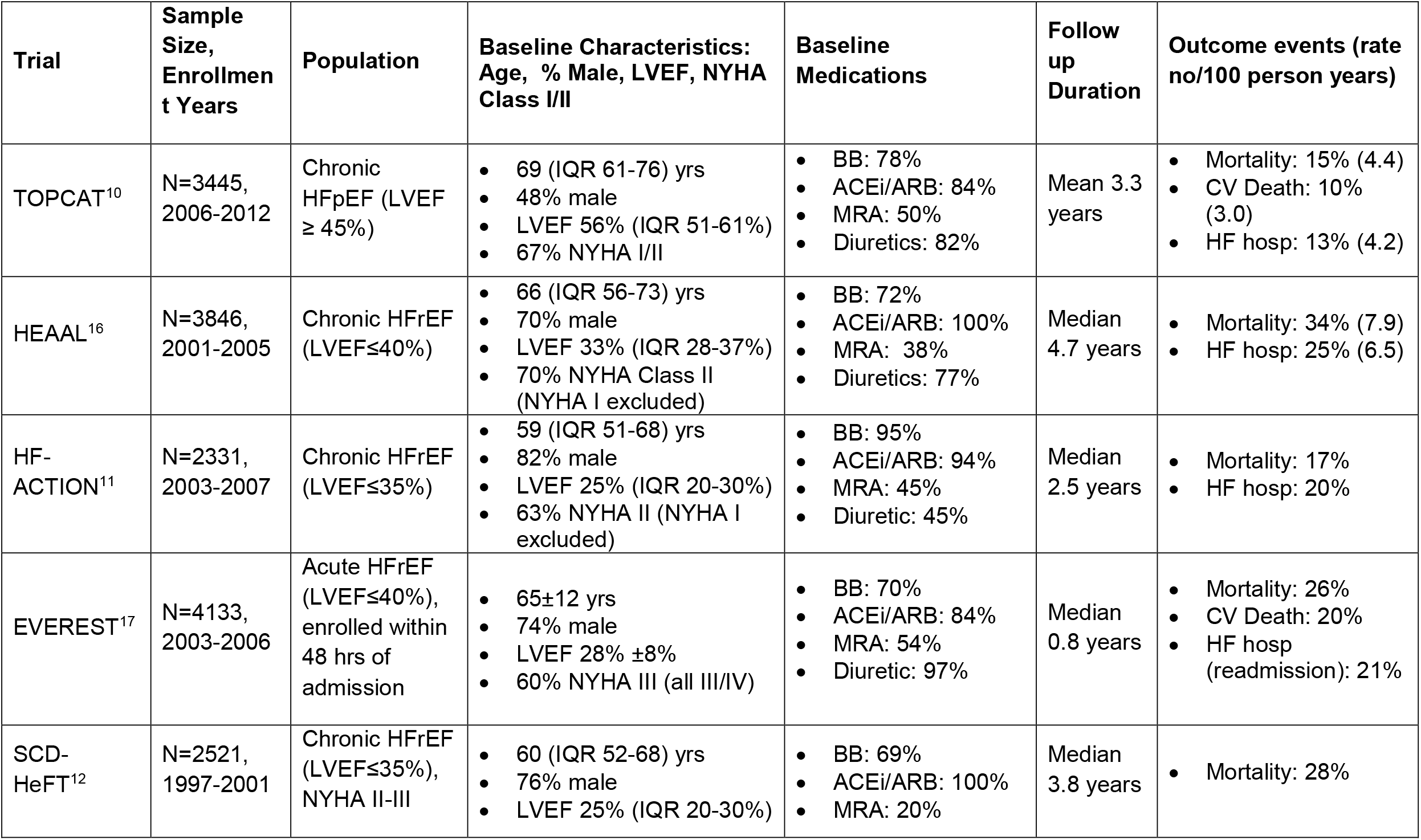
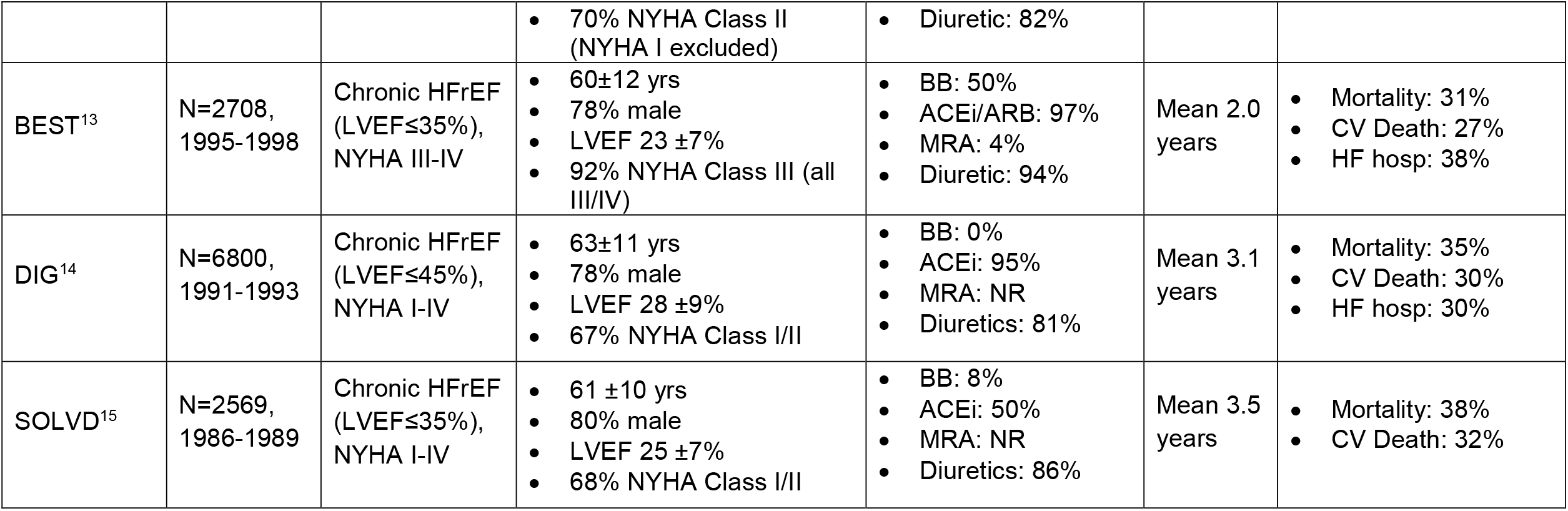
Clinical Trial Databases Used for External Validation

CPM-Dataset Matching: CPMs were matched to validation datasets if the CPM outcomes and variables were available in the validation dataset. We required all CPM variables to be collected in the validation datasets and used a two-step approach in which potential matches were first screened by non-clinical staff for similarity of the index condition and outcomes. This process was then repeated by clinical experts. Finally, CPM-dataset matches were reviewed to identify whether variables and outcomes included in the CPM were available in the validation database. Possible discrepant outcome or variable definitions required review by a physician member of the research team to adjudicate whether the match was appropriate. CPMs were also excluded if a variable was collected in the validation dataset but there was more than 50% missingness.

Observed outcomes in the patient-level data were defined using the CPM outcome definition and prediction time horizon, and observed outcome events that occurred after the prediction time horizon were censored for all model validations. For time-to-event models, Kaplan Meier estimator was used for right-censored follow-up times. For binary outcome models, unobserved outcomes (i.e. due to loss-to-follow-up prior to the prediction time horizon) were considered missing and excluded from analyses.

Relatedness Determination: While all matches between a model and a clinical trial database were deemed to be clinically appropriate, derivation and validations cohorts can be closely or more distantly related in ways that impact model performance. Therefore, a relatedness rubric was developed to characterize the extent to which the CPM derivation cohort and the validation cohorts included similar populations (Supplemental Figure 1). This rubric contained the following categories: 1) Population (chronic HF, acute decompensated HF and NYHA Class); 2) LVEF subtype (reduced LVEF and preserved LVEF); 3) Space/Time (Continent, enrollment years). In addition, for the CPM-dataset matches that included acute HF populations, we included 4) Admission to enrollment timing (within 24 hours of admission, after 24 hours but prior to discharge, at the time of discharge). A HF cardiologist (JNU) compared the characteristics for each derivation-validation dataset pair to determine the relatedness using the following definitions: Closely related was defined as CPMs and datasets with >90% match for all categories, Related as a >90% match for Population and LVEF subtype and 50%-90% match for other categories and Distantly Related as a partial match (>0% but <90%) on at least Population and LVEF subtype. Derivation-validation dataset pairs with no match on Population and LVEF subtype were determined to be inappropriate validations and were excluded.

**Figure 1:**
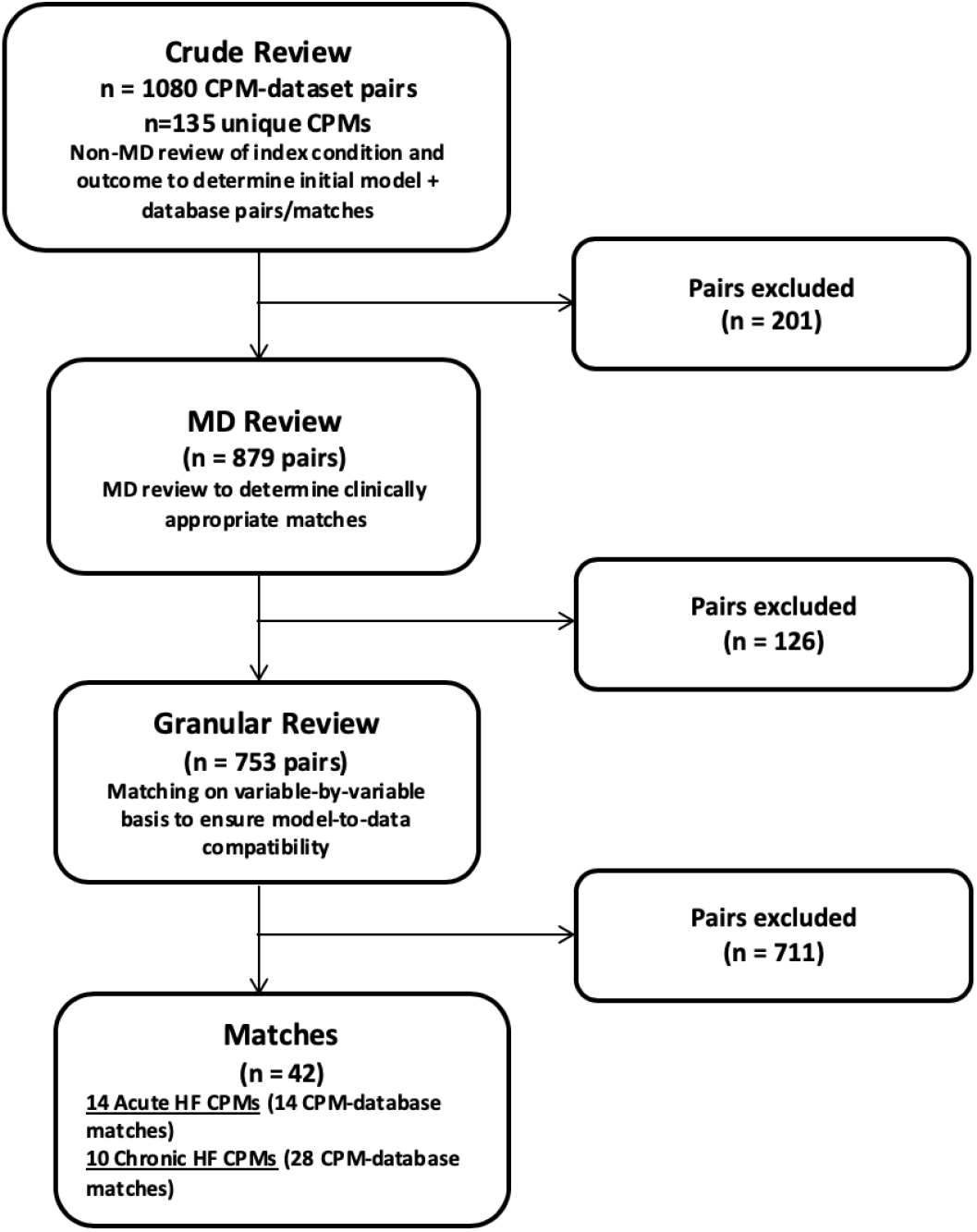

### Measuring CPM Performance

*CPM Discrimination* was evaluated using the c-statistic. The linear predictor for each model was calculated using the published regression coefficients, intercept and the patient-level data from the validation dataset. If only a risk score was reported then this was used for discrimination assessment. Databases with required variables with >50% missingness were excluded. Multiple imputation was performed to impute missing observations in the case of <50% missingness exploiting the correlations between predictor variables and outcomes, respecting the principle of congeniality. Since the c-statistic ranges from 0.5-1.0, the percent change in discrimination was calculated as [(validation c-statistic −0.5) – (Derivation c-statistic −0.5)/(Derivation c-statistic −0.5)*100] to more accurately represent clinically relevant changes. We also calculated the validation database-derived model-based concordance (MB-c) for each validation. The MB-c represents the c-statistic that would result in the validation database if the prediction model were perfectly valid.^18^ Consequently, a difference between the derivation c-statistic and the MB-c is fully due to a difference in case-mix between the derivation and the validation database. The MB-c thus provides a useful benchmark to understand how much the observed changes in discrimination can be attributed to changes in case mix heterogeneity vs model validity.

*CPM Calibration* was assessed by transforming the linear predictor to an event probability. Calibration slope and Harrell’s E statistic standardized to the outcome rate were assessed. Harrell’s E_ave_ computes the average absolute calibration error (the average absolute distance between “observed” outcome rates, based on a smoothed LOESS function and predicted probabilities for each individual); the E_90_ quantifies the 90^th^ percentile of this absolute error.^19^

### Decision curve analysis

Decision curve analyses integrate model discrimination, calibration and the relative utility weights of false-positive and false-negative predictions to present a comprehensive evaluation of the potential clinical consequences of using CPMs to inform treatment decisions across all decision thresholds. For each model, we used decision curve analysis to assess net benefit at 3 decision thresholds: 1) one half of the outcome prevalence rate, 2) the outcome prevalence rate and 3) twice the outcome prevalence.^20^ We standardized net benefit by outcome rate to allow summarizing results across validations. At each decision threshold, the percentage of models with a standardized net benefit above the default strategy (beneficial), equal to the default strategy (neutral) or below the default strategy (harmful) was determined.

### CPM Recalibration

Each CPM-dataset validation underwent three recalibration methods to determine the effect of different model updating procedures. First, the difference between the mean observed outcome rates in the derivation and validation cohorts was used to update the model intercept (calibration-in-the-large). Second, both the intercept and slope were updated to correct for calibration-in-the large (intercept) and to correct for model overfitting through a uniform correction factor for the regression coefficients (slope). Lastly, we re-estimated the regression coefficients using the validation database. All performance measures were repeated on the updated models.

## Results

### CPM-Dataset Matches

1080 potential HF CPM-dataset matches that included 135 individual CPMs were identified. After excluding pairs with no match on population, predictor variables or outcomes, 42 clinically appropriate CPM-dataset matches remained (Figure). These 42 CPM-dataset matches included 24 unique CPMs—14 CPMs derived in a population admitted to the hospital with acute decompensated HF and 10 CPMs derived in outpatients with chronic HF.

### Acute HF CPM Matches

Fourteen CPMs derived in individuals hospitalized with acute HF were able to be matched with the one acute HF validation database, the Effects of oral tolvaptan in patients hospitalized for worsening heart failure: the EVEREST Outcome Trial^17^. The EVEREST trial included 4133 individuals with acute on chronic HF with LVEF of 40% or less, enrolled within 48 hours of admission, randomized to tolvaptan or placebo, followed for a median of 0.8 years with a total of 1080 deaths (26%) (Table 1). The 14 CPMs matched to EVEREST are described in detail in the Supplemental Material (Supplemental Table 1), including population characteristics, variable selection methods, variables included in each CPM, events per variable and discrimination and calibration results in the EVEREST dataset. Of note, one publication reported two different CPMs using the same derivation cohort but different methodology – classification and regression tree analysis and logistic regression^21^. The derivation cohorts were large national multicenter registries for 7 CPMs^21-26^, clinical trials for 3 CPMs^27-29^, single center observational studies for 3 CPMs^30-32^ and an individual patient data meta-analysis (IPDMA) of 30 cohorts for 1 CPM^33^. Two of the 14 CPMs were developed on cohorts of acute HF with reduced LVEF ^26, 27^ while the other 12 CPM derivation cohorts included individuals regardless of LVEF. Similarly, two of the 14 CPMs were developed in a cohort that included both acute and chronic HF^29, 33^, while the remaining 12 on cohorts including only those with acute HF. All CPM-dataset matches except for one were determined to be distantly related; in most cases, this was because the derivation cohort including participants regardless of LVEF, while the validation cohort, EVEREST, only enrolled individuals with reduced LVEF. The outcome for all matched CPMs was all-cause mortality. The CPM mortality outcome assessment was in-hospital in 5 CPMs^21, 22, 24, 31^, 60 days for 1 CPM^27^, 60-90 days for 1 CPM^23^, 12 weeks for 1 CPM^25^, 6 months for 1 CPM^28^, 1 year for 4 CPMs^26, 30, 32, 33^ and 18 months for 1 CPM^29^. In each case, we used the CPM-defined outcome timeframe for external validation. The majority of CPMs used logistic regression with stepwise variable selection while 1 CPM used classification and regression tree analysis. The number of variables in the final models ranged from 3 to 13 with 8 out of 14 models having 5 or fewer variables. Commonly included variables in the CPMs included renal function (14 out of 14 CPMs), age (10 CPMs), systolic blood pressure (9 CPMs), serum sodium (8 CPMs), LVEF (5 CPMs), lung disease (5 CPMs) and heart rate (4 CPMs). Events (in the respective derivation cohorts) per variable in the final models ranged from 6 to 346 with 5 models having less than 20 events per variable and 2 models having less than 10 events per variable. Review of published literature through 2015 identified that 8 (57%) of the acute HF CPMs had been externally validated at least once.

### Chronic HF models

Ten CPMs derived in cohorts with chronic HF were matched to the validation datasets that included individuals with chronic ambulatory HF, yielding a total of 28 unique CPM-dataset matches. The validation datasets are described in detail in Table 1 and the CPMs are described in Supplemental Table 2. Of the 10 CPMs, 1 CPM was matched to 5 validation datasets, 3 CPMs were matched to 4 validation datasets, 2 CPMs were matched to 3 validation datasets, 1 CPM was matched to 2 validation datasets and 3 CPMs were matched to 1 validation dataset. Derivation cohorts for matched CPMs were single center observational studies for 5 CPMs,^34-37^ clinical trial cohorts for 3 CPMs,^29, 38, 39^ multicenter registry for 1 CPM,^40^ and an observational study from 2 centers for 1 CPM.^41^ Two CPMs were derived in cohorts of HF regardless of LVEF,^29, 37^ while the other 8 models included a population with reduced LVEF. No matched CPMs were derived in a population with HF with preserved LVEF. One CPM derivation cohort included individuals with both acute and chronic HF,^29^ while the other 9 models were derived in cohorts of individuals only with chronic HF. Seven CPMs were derived in populations defined by primary prevention implantable cardioverter-defibrillator (ICD) implantation.^34-36, 39-41^ The CPM outcome was CV mortality for 1 CPM,^37^ death without prior ICD therapy for 1 CPM,^35^ and all-cause mortality for 8 CPMs, with the same outcome used in matched validation cohorts. All models used regression equations with most utilizing stepwise selection techniques. The number of variables in the final models ranged from 4 to 8 with 7 out of 10 models having 5 or fewer variables. Events in the derivation cohort per variable included in the CPM ranged from 6 to 963 with only 1 CPM having less than 10 events per variable. Commonly included variables in the CPMs included age (9 out of 10 CPMs), renal function (8 CPMs), LVEF (5 CPMs), diabetes (5 CPMs), atrial fibrillation (4 CPMs) and NYHA Class (4 CPMs). Review of published literature through 2015 identified that 3 (30%) of the chronic HF CPMs had been externally validated at least once.

**Table 2.** Validation Performance Acute CHF All Matches

| (n = 14) | Mean (SD) | Median (IQR) | Range |
| --- | --- | --- | --- |
| *Discrimination* |  |  |  |
| Development c-statistic | 0.77 (0.05) | 0.76 (0.75, 0.8) | 0.69, 0.86 |
| Validation c-statistic | 0.67 (0.03) | 0.67 (0.65, 0.68) | 0.62, 0.73 |
| Validation model-based c-statistic (MBc) | 0.71 (0.07) | 0.68 (0.66, 0.76) | 0.61, 0.84 |
| *% Change in discrimination due to...* |  |  |  |
| Total (val.c vs. dev.c) | -37 (15) | -40 (-49, -19) | -55, -11 |
| Case mix heterogeneity (MBc vs. dev.c) | -20 (19) | -24 (-33, -5) | -57, 8 |
| Model validity (val.c vs. MBc) | -12 (24) | -8 (-31, 5) | -53, 27 |
| **Calibration** |  |  |  |
| Slope | 0.8 (0.26) | 0.87 (0.57, 1) | 0.34, 1.15 |
| *standardized E* | 1.5 (2) | 0.5 (0.4, 2.2) | 0.1, 7.1 |
| *standardized E90* | 2.8 (3.6) | 1.1 (0.5, 3.4) | 0.2, 12.9 |

### Acute HF CPM Validation Results

#### Discrimination

Derivation c-statistics were reported in 10 out of 14 CPMs and ranged from 0.69 to 0.86 (median 0.76, IQR 0.75, 0.8) (Table 2). Observed validation c-statistic ranged from 0.62 to 0.73 (median 0.67, IQR 0.65, 0.68) and model-based c-statistic ranged from 0.61 to 0.84 (median 0.68, IQR 0.66, 0.76). For the 10 CPMs that reported a derivation c-statistic, the median percent decrement between the derivation and validation cohort c-statistic was −40% (IQR −49%, −19%) (Table 2). For most models, the decrement in discrimination was largely due to less case-mix heterogeneity in the EVEREST cohort (median percent change between derivation and model-based c-statistic was −24%, IQR −33%, −5%) and to a lesser extent to model validity (the median percent decrement between the model-based c-statistic and the validation cohort was −8%, IQR –31%, 5%). CPMs developed in smaller cohorts tended to have the greatest decrement in discrimination between both validation and derivation as well as validation and model-based c-statistic, suggesting that the decrement was mostly due to model validity and not case-mix (Supplemental Table 1).

#### Calibration

The median calibration slope for the 14 CPM models was 0.87 (IQR 0.57, 1.0), the median E_avg_ standardized to the outcome rate was 0.5 (IQR 0.4, 2.2) and the median standardized E_90_ was 1.1 (IQR 0.5, 3.4) (Table 2). Calibration was worse for the 4 models predicting in-hospital mortality with E_avg_ ranging from 1.44-7.1 in these models (Supplemental Table 1). Calibration was better for models predicting outcomes over a longer time frame from 3 months to 18 months with standardized E for these 9 models ranging from 0.06-0.6.

#### Decision curve analysis

At a threshold set to one half the outcome rate, 29% of CPMs were harmful, 14% were beneficial and 57% would yield outcomes similar to the default strategy (neutral) (Table 3). Using a threshold set to the outcome rate, 29% of CPMs were harmful and 71% of CPMs were beneficial. At a threshold of twice the outcome rate, 57% were harmful and 29% were beneficial. CPMs predicting in-hospital mortality suggested negative or marginal net benefit at the observed outcome prevalence (indicating these models would be useless at best in the EVEREST cohort), while models predicting longer term outcomes (with higher event rates) showed positive net benefit at the observed outcome prevalence with variable benefit at lower and higher prevalence thresholds (Supplemental Table 1).

**Table 3.** Net Benefit Compared to Default Strategy Acute CHF All Matches

| Validation | Threshold | Compared to default strategy |  |  |
| --- | --- | --- | --- | --- |
|  |  | % Above | % Neutral* | % Below |
| Original model | Prev./2 | 14.3 | 57.1 | 28.6 |
|  | Prevalence | 71.4 | 0.0 | 28.6 |
|  | Prev.*2 | 28.6 | 14.3 | 57.1 |
| Updated intercept | Prev./2 | 42.9 | 42.9 | 14.3 |
|  | Prevalence | 100.0 | 0.0 | 0.0 |
|  | Prev.*2 | 64.3 | 14.3 | 21.4 |
| Updated intercept and slope | Prev./2 | 42.9 | 50.0 | 7.1 |
|  | Prevalence | 100.0 | 0.0 | 0.0 |
|  | Prev.*2 | 64.3 | 21.4 | 14.3 |
| Re-estimated | Prev./2 | 35.7 | 42.9 | 21.4 |
|  | Prevalence | 100.0 | 0.0 | 0.0 |
|  | Prev.*2 | 78.6 | 7.1 | 14.3 |

\* Neutral is defined as NB equal to the default strategy

#### Model updating

For acute HF CPMs, most of the calibration error was corrected by updating the model intercept (median % change in E −81%, IQR −93%, −19%), with slight incremental improvement in calibration with updating both the intercept and the slope (incremental median % change in E −22%, IQR −73%, 0%) and from re-estimating the model (incremental median % change in E −13%, IQR −79%, 50%) (Table 4). The net benefit of using a CPM improved with updating the model intercept alone: 100% of CPMs were beneficial at the outcome prevalence and a smaller percentage of CPMs were harmful at the other thresholds (Tables 3 and 4).

**Table 4.** Effects of Updating Acute CHF All Matches

| (n = 14) | Intercept | Intercept + Slope |  | Re-estimation |  |
| --- | --- | --- | --- | --- | --- |
|  | *Compared to Original* | *Compared to Original* | *Incremental Change* | *Compared to Original* | *Incremental Change* |
| **% Change in Discrimination** |  |  |  |  |  |
| c-statistic | N/A | N/A | N/A | 9 (5, 13) | N/A |
| MB c-statistic | N/A | N/A | N/A | 1 (-25, 15) | N/A |
| **% Change in Calibration Error** |  |  |  |  |  |
| E | -81 (-93, -19) | -91 (-93, -80) | -22 (-73, 0) | -95 (-97, -85) | -13 (-79, 50) |
| E90 | -84 (-94, -47) | -88 (-94, -77) | -22 (-56, 17) | -94 (-98, -87) | -17 (-43, 0) |
| **% Change in Standardized Net Benefit** |  |  |  |  |  |
| sNB prev. / 2 | 7 (-1, 119) | 97 (2, 119) | 0 (0, 49) | 76 (13, 97) | 11 (-49, 46) |
| sNB prev. | 13 (0, 229) | 13 (0, 229) | 0 (0, 0) | 31 (11, 277) | 14 (11, 33) |
| sNB prev. * 2 | 109 (39, 163) | 109 (95, 163) | 0 (0, 4) | 169 (113, 289) | 36 (4, 82) |

\*All values are reported as median (IQR).

### Chronic HF CPM Validation Results

#### Discrimination

Derivation c-statistics were reported in 8 out of 10 CPMs and ranged from 0.73 to 0.81 (median 0.76, IQR 0.74, 0.8) (Table 5). Observed validation c-statistics ranged from 0.53 to 0.70 (median 0.61, IQR 0.6, 0.63) and the model-based c-statistics ranged from 0.53 to 0.78 (median 0.68, IQR 0.62, 0.71). For the 8 CPMs that reported a derivation c-statistic, the median percent decrement between the derivation and validation cohort c-statistic was −55% (IQR −62%, −48%). Unlike the Acute HF CPMs, for most Chronic HF CPMs the decrement in discrimination appeared to be due to both case-mix heterogeneity (median percent change between derivation and model-based c-statistic was −28%, IQR −36%, −12%) and model invalidity (median percent decrement between the model-based c-statistic and the validation cohort was −34%, IQR –48%, 0%) (Table 5).

**Table 5.** Validation Performance Chronic CHF All Matches

| (n = 28) | Mean (SD) | Median (IQR) | Range |
| --- | --- | --- | --- |
| **Discrimination** |  |  |  |
| Development c-statistic | 0.77 (0.03) | 0.76 (0.74, 0.8) | 0.73, 0.81 |
| Validation c-statistic | 0.61 (0.04) | 0.61 (0.6, 0.63) | 0.53, 0.7 |
| Validation model-based c-statistic (MBc) | 0.67 (0.07) | 0.68 (0.62, 0.71) | 0.53, 0.78 |

| *% Change in discrimination due to...* |  |  |  |
| --- | --- | --- | --- |
| Total (val.c vs. dev.c) | -54 (16) | -55 (-62, -48) | -86, -22 |
| Case mix heterogeneity (MBc vs. dev.c) | -24 (13) | -28 (-36, -12) | -44, -7 |
| Model validity (val.c vs. MBc) | -13 (65) | -34 (-48, 0) | -78, 234 |
| *Calibration* |  |  |  |
| Slope | 0.52 (0.31) | 0.46 (0.33, 0.58) | 0.17, 1.56 |
| standardized E | 0.6 (0.5) | 0.5 (0.3, 0.7) | 0.2, 2.9 |
| standardized E90 | 1.1 (1.5) | 0.7 (0.5, 1) | 0.2, 6.8 |

#### Calibration

The median calibration slope for the 10 CPMs was 0.46 (IQR 0.33, 0.58), the median E_avg_ standardized to the outcome rate was 0.5 (0.3, 0.7) and the median standardized E_90_ was 0.7 (0.5, 1.0) (Table 5). Five out of 28 CPM-dataset matches were related and the rest were distantly related. In the CPMs with related and distantly related matches, there was no signal for better model performance in related CPM-dataset matches. In CPMs with multiple database matches, there was a range of discrimination and calibration results and no CPMs had consistently good discrimination and calibration across validation samples (Supplemental table 3).

#### Decision curve analysis

At a threshold set to one half the outcome rate, 64% of chronic HF CPMs were harmful, 7% were beneficial and 29% would yield outcomes similar to the default strategy (neutral) (Table 6). Using a threshold set to the outcome rate, 4% of CPMs were harmful and 86% of CPMs were beneficial. At a threshold of twice the outcome rate, 43% were harmful, 14% were beneficial and 43% were neutral.

**Table 6.**
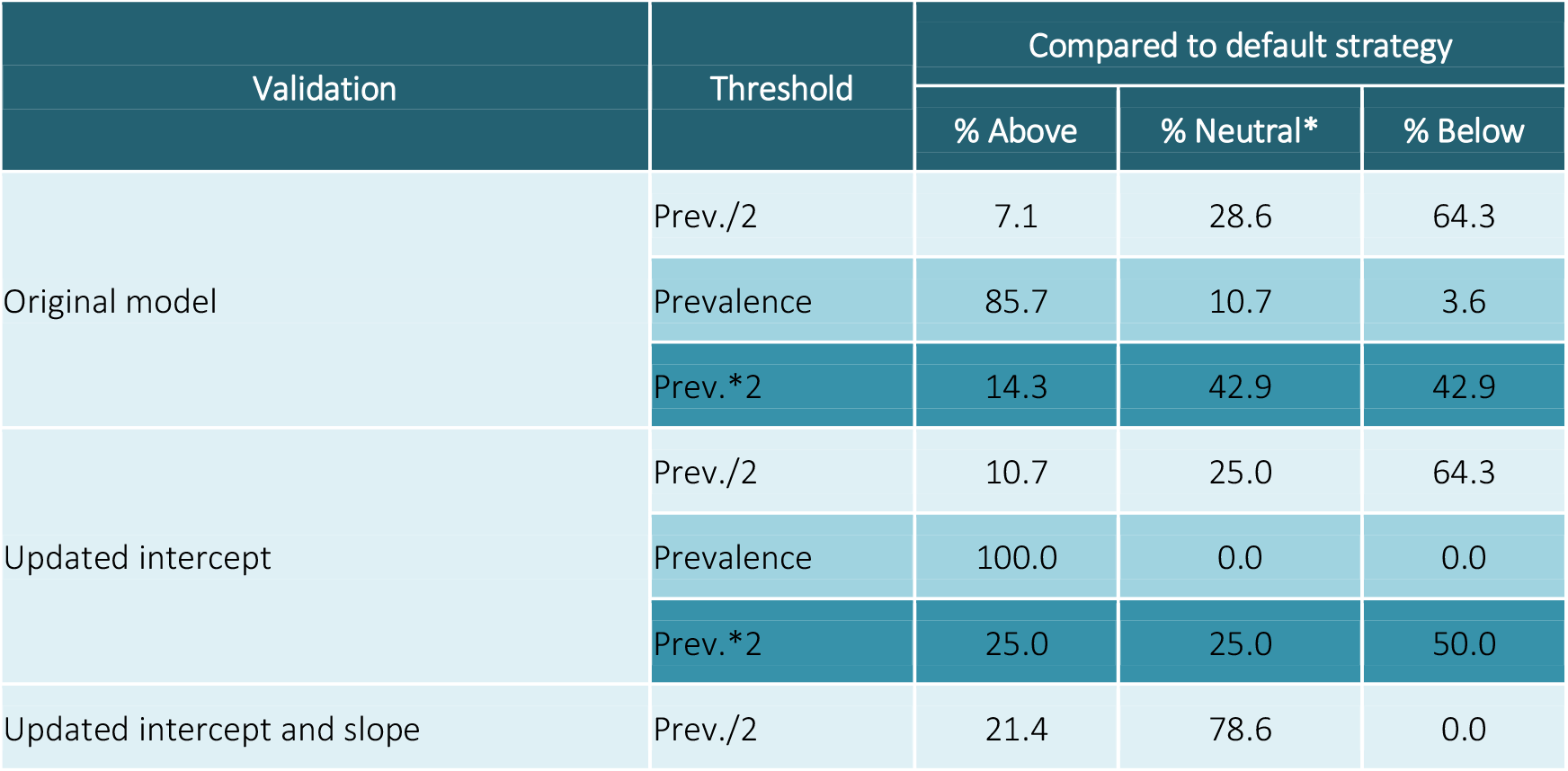
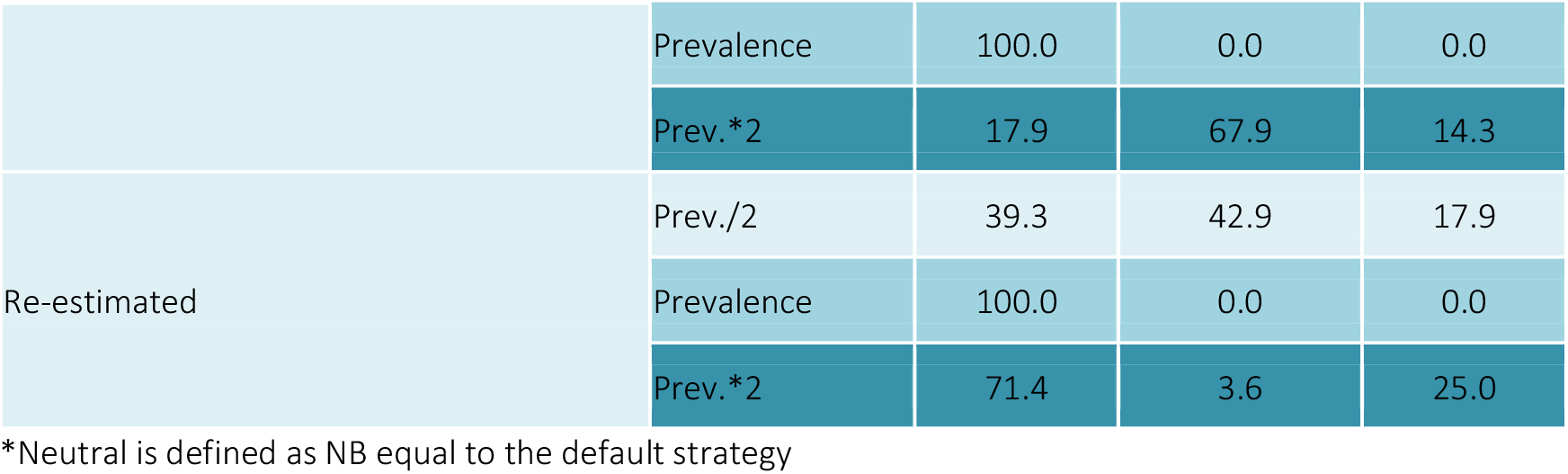
Net Benefit Compared to Default Strategy Chronic CHF All Matches

#### Model updating

Unlike with the acute HF CPMs, updating the model intercept only slightly improved calibration (median % change in E_avg_ −23%, IQR −54%, 0%); however, updating both the intercept and the slope led to incremental improvement in calibration (incremental median % change in E_avg_ −94%, IQR −99%, −80%) (Table 7). There was marginal improvement in calibration from re-estimating the model (incremental median % change in E_avg_ 0% (IQR −50%, 0%). The net benefit of using a CPM improved with model updating but updating of both the intercept and the slope was required to prevent net harm at one half the outcome prevalence and twice the outcome prevalence (Tables 6 and 7).

**Table 7.** Effects of Updating Chronic CHF All Matches

| (n = 28) | Intercept | Intercept + Slope |  | Re-estimation |  |
| --- | --- | --- | --- | --- | --- |
|  | *Compared to Original* | *Compared to Original* | *Incremental Change* | *Compared to Original* | *Incremental Change* |
| **% Change in Discrimination** |  |  |  |  |  |
| c-statistic | N/A | N/A | N/A | 15 (6, 30) | N/A |
| MB c-statistic | N/A | N/A | N/A | -29 (-42, 1) | N/A |
| **% Change in Calibration Error** |  |  |  |  |  |
| E | -23 (-54, 0) | -96 (-100, -84) | -94 (-99, -80) | -98 (-100, -92) | 0 (-50, 0) |
| E90 | -26 (-64, 17) | -98 (-100, -89) | -95 (-100, -76) | -98 (-99, -90) | 0 (-27, 20) |
| **% Change in Standardized Net Benefit** |  |  |  |  |  |
| sNB prev. / 2 | 0 (0, 61) | 100 (100, 100) | 100 (100, 100) | 100 (100, 138) | 84 (-7, 169) |
| sNB prev. | 0 (0, 36) | 0 (0, 71) | 0 (0, 0) | 41 (19, 97) | 21 (9, 49) |
| sNB prev. * 2 | 38 (0, 100) | 100 (0, 100) | 100 (0, 100) | 108 (35, 149) | 55 (-16, 318) |

\*All values are reported as median (IQR).

## Discussion

We report the first systematic approach to HF CPM validation, including a systematic review to identify published HF CPMs, matching of model variables and outcomes with available clinical trial datasets and performance of CPM external validation of all CPMs with a matched dataset. We report discrimination, calibration, net benefit and the effect of model updating on predictive performance. Our principal findings are as follows: 1) using “off the shelf” CPMs may cause net harm unless the CPM is well calibrated in a given clinical population; 2) a minority of published CPMs were compatible with available datasets and even when compatible were often derived on datasets that were “distantly related” to the validation data; 3) less case mix heterogeneity, estimated using the model-based c-statistic, explained most of the decrement in discrimination in the matched acute HF CPMs but not the chronic HF CPMs.

The acute HF CPMs that were able to be matched to the EVEREST clinical trial database had largely preserved discrimination when accounting for case-mix and our analysis suggests that many of these models could be appropriate for clinical care. Calibration was suboptimal in the acute HF CPM validations but this was likely due to the differences in derivation and validation cohorts and calibration measures and net benefit were improved with updating of the model intercept (calibration-in-the-large). The chronic HF CPMs that were matched to clinical trial databases were largely derived from ICD cohorts and had a significant decrement in discrimination that was only partially explained by case-mix. Calibration was also suboptimal for chronic HF CPMs and required updating of both the intercept and slope to improve calibration and net benefit.

The MB-c statistic allows us to determine how much of a change in discrimination is due to differences in case mix between the development and validation cohort rather than CPM validity. This is an important distinction when interpreting changes in discrimination in validation settings and the MB-c allows for a more nuanced understanding of discrimination in external validation studies. The validation datasets represented clinical trial cohorts while many of the CPMs were derived in registry cohorts, thus it is not surprising that the MB-c, and consequently the observed c-statistic was lower than the derivation c-statistic in this setting. Applying the MB-c in external validation studies should be routinely performed.

Our findings demonstrate the potential for harm by using CPMs with poor discrimination or that are poorly calibrated in the population in which they are applied. Decision curve analyses integrate model performance (including both discrimination and calibration) and the relative utility weights of false-positive and false-negative predictions across all decision thresholds to present a comprehensive evaluation of the potential clinical consequences of using CPMs to inform treatment decisions. Our results suggest that using a CPM for decision making when the threshold is far from the average (e.g. less than half or greater than twice the average outcome incidence) is not recommended unless one knows that the CPM is well calibrated in your clinical population. For the acute HF models, updating the model intercept (i.e. calibration in the large) improved the net benefit substantially, suggesting that simple model updating can substantially improve CPM performance and clinical decision making.

There are limitations to our analysis that should be considered. The validation databases comprised clinical trials and thus represent a more selected clinical population than the registries or real-world populations from which many HF CPMs were derived. However, our inclusion of the MBc allows us to assess the contribution of case-mix to decrements in discrimination between derivation and validation cohorts. Some of the validation and CPM cohorts enrolled participants in older treatment eras; however, these external validations still demonstrate the approach that can be used to evaluate CPM performance and recalibrate CPMs. We were only able to match a minority of the CPMs with one of the 8 databases and thus were not able to evaluate the performance of many published CPMs.

In conclusion, using “off the shelf” CPMs may cause net harm unless the CPM is well calibrated in a given clinical population or the CPM is recalibrated in a population similar to the one in which the model will be applied in practice. We were only able to externally validate a minority of CPMs due to mismatch in predictor variables, outcomes or population between the CPMs and the validation dataset, suggesting that CPMs often use variables not routinely collected. When CPMs were able to be matched, there was often a decrement in discrimination and poor calibration. Our findings demonstrate the importance of applying underutilized techniques, including both the model-based c-statistic and decision curve analysis. These approaches showed that a large proportion of the decrement in discrimination was due to changes in the case mix between the derivation and validation populations, and that the high risk of net harm of using off the shelf models can be substantially mitigated by simple model recalibrations. How to achieve recalibration in actual clinical practice is important if CPMs are to realize their potential for improving decision making and health outcomes in routine care.

## Supporting information

PRISMA checklist

Supplemental File Search Terms

## Data Availability

Tufts Predictive Analytics and Comparative Effectiveness (PACE) CPM Registry

## Acknowledgement

Research reported in this work was funded through a Patient-Centered Outcomes Research Institute® (PCORI®) Award (ME-1606-35555).

