## Supplemental File Search Terms for "Performance of Heart Failure Clinical Prediction Models: A Systematic External Validation Study"

Short title: HF prediction model external validation

Keywords: Clinical prediction model, heart failure, mortality

Acknowledgement: Research reported in this work was funded through a Patient-Centered Outcomes Research Institute® (PCORI®) Award (ME-1606-35555).

Disclaimer: The views, statements, opinions presented in this work are solely the responsibility of the author(s) and do not necessarily represent the views of  the Patient-Centered Outcomes Research Institute® (PCORI®), its Board of Governors or Methodology Committee.

Supplemental Material

Systematic Review Search Terms:

((predict$ adj1 model$) or (predict$ adj1 instrument$) or (predict$ adj1 score$) or (predict$ adj1

index)).mp.

((prognos$ adj1 model$) or (prognos$ adj1 instrument$) or (prognos$ adj1 score$) or (prognos$ adj1

index)).mp.

((risk adj1 model$) or (risk adj1 instrument$) or (risk adj1 score$) or (risk adj1 index) or (risk

assessment model or risk assessment instrument or risk assessment score)).mp.

atrial fib$.mp. or exp Atrial Fibrillation/ or exp coronary artery disease/ or exp coronary disease/ or exp

myocardial infarction/ or Myocardial infarct$.mp. or exp Heart Failure, Congestive/ or exp myocardial

ischemia/ or exp cardiovascular diseases/ or exp Cerebrovascular Accident/ or *heart failure/ or *stroke/

or *acute coronary syndrome/

limit 6 to yr="1990 -Current"

**Where current = May 15, 2015 publications

(201205$ or 201206$ or 201207$ or 201208$ or 201209$ or 201210$ or 201211$ or 201212$ or 2013$

or 2014$ or 201501$ or 201502$ or 201503$).ed.
