## Supplemental Table 1 for "Performance of Heart Failure Clinical Prediction Models: A Systematic External Validation Study"

Short title: HF prediction model external validation

Keywords: Clinical prediction model, heart failure, mortality

Acknowledgement: Research reported in this work was funded through a Patient-Centered Outcomes Research Institute® (PCORI®) Award (ME-1606-35555).

Disclaimer: The views, statements, opinions presented in this work are solely the responsibility of the author(s) and do not necessarily represent the views of  the Patient-Centered Outcomes Research Institute® (PCORI®), its Board of Governors or Methodology Committee.

Supplemental Table 1: Acute HF Clinical Prediction Models with EVEREST Database Matches:

| CPM Citation | Sample Size, Years enrollment | Population (relatedness to EVEREST validation cohort) | Baseline Characteristics:  Age, % Male, LVEF | Variable Selection | Variables final model | Events per variable (EPV) in Derivation Cohort | CPM Outcome Definition | Outcome, N(%) | Derivation c-statistic and Calibration (if reported) | Validation Results: Discrimination in the EVEREST Cohort | Validation Results: Calibration in the EVEREST Cohort: *E and E90 standardized to outcome rate | Number of prior validations reported in literature (through 2015) |
| --- | --- | --- | --- | --- | --- | --- | --- | --- | --- | --- | --- | --- |
| Felker et al^27^ | N=949, 1997-1999 | Acute HFrEF, OPTIME-CHF trial (Related) | - 68 (56-76) yrs - 66% male - LVEF 23% (18-30%) | Backward stepwise selection | - Age - SBP - BUN - Sodium - NYHA Class | 18 | Mortality at 60 days (includes in-hospital mortality | Derivation Cohort: N=90 (9.6%)  Validation Cohort: N=283 (7.2%) | C-statistic: 0.77 | C-statistic: 0.73  % change in C-statistic (Val vs. Der):-16%  Model Based C statistic: 0.76  % change in C-statistic (Val vs MBC):-11% | Calibration slope: 0.81  E: 4.2%  E*: 0.58  E90: 10.9%  E90*: 1.51  Net Benefit (prev): 0.29 | 0 |
| Peterson et al^22^ | N=39,783; 2005-2007 | Acute HF, all LVEF, GWTG-HF  (Related) | - 73 (IQR 59-87) yrs - 49% male - LVEF 39% (IQR 22-55%) | Variables p<0.05 in multivariable analysis included | - Age - SBP - BUN - Sodium - Heart Rate - Race - COPD | 162 | In-hospital mortality | Derivation Cohort: N=1139 (3%)  Validation Cohort: N=36 (0.9%) | C-statistic: 0.75;  Hosmer-Lemeshow (p=0.888) | C-statistic: 0.65  % change in C-statistic (Val vs. Der):-39%  Model Based C statistic: 0.67  % change in C-statistic (Val vs MBC):-8.9% | Calibration slope: 1.11  E: 2%  E*: 2.2  E90: 3%  E90*: 3.3  Net Benefit (prev): -0.12 | 2 |
| O’Connor et al^23^ | N=4,402, 2003-2004 | Acute HF, all LVEF, OPTIMIZE-HF registry (10% subset with postdischarge data)  (Distantly Related) | - 72±14 yrs - 51% male - LVEF 37%±17% | Cox PH model with 19 prespecified variables with forward stepwise and backward variable selection | - Age - SBP - Creatinine - Sodium - COPD/asthma - Liver disease - Admit weight - Depression | 60 | Mortality at 60 days post discharge | Derivation Cohort: N=481 (8.6%)  Validation Cohort: N=169 (7%) | C-statistic: 0.72  Calibration plot (deciles) | C-statistic: 0.68  C-statistic (Val vs. Der):-18.6%  Model Based C statistic: 0.66  % change in C-statistic (Val vs MBC):14% | Calibration slope: 1.15  E: 3%  E*: 0.43  E90: 6.8%  E90*: 0.97  Net Benefit (prev):0.23 | 2 |
| Abraham et al^24^ | N=37,548 2003-2004 | Acute HF, all LVEF, OPTIMIZE-HF registry  (Distantly Related) | - 73±14 yrs - 48% male - LVEF 39%±18% | Logistic regression45 candidate variables. Stepwise and backward variable selection (complete case ) with p<0.05. | - Age - SBP - Creatinine - Sodium - Heart rate - HF primary cause of admission - LV systolic dysfunction | 262 | In-hospital mortality | N=1,834 (3.8%)  Validation Cohort: 36 (0.9%) | C-statistic:0.75 | C-statistic: 0.67  C-statistic (Val vs. Der):-31.6%  Model Based C statistic: 0.66  % change in C-statistic (Val vs MBC): 5.5% | Calibration slope: 1.03  E: 1.3%  E*: 1.44  E90: 3.1%  E90*: 3.44  Net Benefit (prev): -0.13 | 2 |
| Fonarow et al^21^  CART model | N=33,046, 2001-2003 | Acute HF, all LVEF,  ADHERE HF registry  (Distantly Related) | - 73±14 yrs - 48% male - 56% LVEF <40% | Classification and Regression Tree Analysis (CART) | - SBP - BUN - Creatinine | 461 | In-hospital mortality | N=1383 (4.2%)  Validation Cohort: 36 (0.9%) | C-statistic: 0.69 | C-statistic: 0.67  C-statistic (Val vs. Der): -10.7%  Model Based C statistic: 0.68  % change in C-statistic (Val vs MBC): -6.7% | Calibration slope: 0.90  E: 3.7%  E*: 4.11  E90: 5.2%  E90*: 5.78  Net Benefit (prev): -0.13 | 5 |
| Fonarow et al^21^  Regression model | N=33,046, 2001-2003 | Acute HF, all LVEF,  ADHERE HF registry  (Distantly Related) | - 73±14 yrs - 48% male - 56% LVEF <40% | Logistic regression | - Age - SBP - BUN - HR | 346 | In-hospital mortality | N=1383 (4.2%)  Validation Cohort: 36 (0.9%) | C-statistic: 0.76 | C-statistic: 0.65  C-statistic (Val vs. Der): -43.6%  Model Based C statistic: 0.693  % change in C-statistic (Val vs MBC): -24.4 | Calibration slope: 0.91  E:2.9%  E*: 3.22  E90:6.1%  E90*: 6.78  Net Benefit (prev): -0.12 | 5 |
| Velavan et al^25^ | N=10,701, 2000-2001 | Acute HF, all LVEF, Euro HF Survey  (Distantly Related) | - 71±13 yrs - 53% male - 35% Moderate-severe LVSD | Stepwise logistic regression | - Age - Cr - LVEF (mild, moderate, severe) - ACEi/ARB use - BB use | 281 | Mortality at 12 weeks (includes in-hospital) | N=1404 (13%)  Validation Cohort: 367 (9.5%) | NR | C-statistic: 0.67  C-statistic (Val vs. Der): NR  Model Based C statistic: 0.67  % change in C-statistic (Val vs MBC): 1.2% | Calibration slope: 1.001  E: 4.2%  E*: 0.44  E90: 6.6%  E90*: 0.69  Net Benefit (prev): 0.25 | 0 |
| Pocock et al^33^ | N=39,372, years not reported | Individual data meta-analysis, 31 studies, all LVEF both acute and chronic HF populations  (Distantly Related) | - 66±11 yrs - 67% male - LVEF 35%±14% - 56% NYHA I/II | Forward stepwise selection using p<0.01, poisson regression | - Age - Gender - BMI - Current smoke - SBP - Diabetes - NYHA Class - LVEF - COPD - HF duration >18 months - Creatinine - Beta-blocker - ACEi/ARB | 1056 | All cause mortality. 1 year and 3 year timepoints reported | Validation Cohort: 822 (34.8 %) | NR | C-statistic: 0.68  C-statistic (Val vs. Der): NR  Model Based C statistic: 0.68  % change in C-statistic (Val vs MBC): 4.3% | Calibration slope: 0.95  E: 1.3%  E*: 0.37  E90: 17.5%  E90*: 0.5  Net Benefit (prev): 0.14  **Above using the 1 year predictions | 1 |
| Scrutinio et al^30^ | N=453, years not reported | Acute HF, all LVEF  (Distantly Related) | - 68±12 yrs - 76% male - LVEF 31%±12% - NYHA Class 3.4±0.6 | Logistic regression including variables with p<0.10 | - LVEF - Hemoglobin - BUN - Sodium - COPD | 25 | All cause Mortality at 1 year | N=128 (28%);  N=954 (26%) | C-statistic: 0.83 | C-statistic: 0.67  C-statistic (Val vs. Der): -48%  Model Based C statistic: 0.75  % change in C-statistic (Val vs MBC): -31% | Calibration slope: 0.57  E: 7%  E*: 0.27  E90: 12.3%  E90*: 0.47  Net Benefit (prev): 0.24 | 0 |
| Kinugasa et al | N=349, 2004-2008 | Acute HF, all LVEF, single center  (Distantly Related) | - 68±12 yrs - 56% male - 34% LVEF <40% | Forward, stepwise multivariate logistic regression p<0.05 | - Previous HF hospitalization - BUN - Sodium - Albumin - BNP | 7 | In-hospital mortality | N=34 (9.7%)  N=31 (1.1%) | C-statistic = 0.86 | C-statistic: 0.72  C-statistic (Val vs. Der): -40%  Model Based C statistic: 0.84  % change in C-statistic (Val vs MBC): -36% | Calibration slope: 0.50  E: 7.8%  E*: 7.09  E90: 14.2%  E90*: 12.9  Net Benefit (prev): 0.07 | 0 |
| Martinez-Selles et al | N=701, 1996 | Acute HF, all LVEF, single center  (Distantly Related) | - 72±12 yrs - 45% male - 34% LVEF <30% | Backward, stepwise Cox PH model | - Age - LVEF - Creatinine - COPD - Stroke - Aortic stenosis | 78 | Mortality or heart transplantation | N=470 (67%) at a median f/u of 5 years  N=958 (26%) at 1 year | C-statistic = 0.76 | C-statistic: 0.62  C-statistic (Val vs. Der): -55%  Model Based C statistic: 0.61  % change in C-statistic (Val vs MBC): 6.3% | Calibration slope: 0.84  E: 3%  E*: 0.12  E90: 5.5%  E90*: 0.21  Net Benefit (prev): 0.19 | 0 |
| Huynh et al | N=288, 1990-1994 | Acute HF, age 70 years and older, all LVEF, single center, enrolled in disease management study  (Distantly Related) | - 79±6 yrs - 37% male - LVEF 42%±12% - NYHA Class 2.3±1 | Multivariate logistic regression, variables with p<0.10 considered | - BUN - SBP - PAD - Sodium | 11 | Mortality at 6 months | N=43 (15%)  311 (22%) | C-statistic = 0.80 | C-statistic: 0.65  C-statistic (Val vs. Der): -51%  Model Based C statistic: 0.79  % change in C-statistic (Val vs MBC): -48% | Calibration slope: 0.43  E: 10%  E*: 0.46  E90: 27%  E90*: 1.2  Net Benefit (prev): 0.24 | 0 |
| Alla et al | N=219, 1994 | Acute HF, LVEF ≤ 30%, community based in France  (Distantly Related) | - Mean age 65 yrs - 78% male - 22% have LVEF <18.5% | Forward stepwise logistic regression, separate models for ischemic and non-ischemic cardiomyopathy | - Age - HR - Creatinine - Sodium - Prior HF hosp | 20 | Mortality at 1 year | N=99 (45%)  683 (27%) | NR | C-statistic: 0.64  Model Based C statistic: 0.61  % change in C-statistic (Val vs MBC): +27% | Calibration slope: 0.73  E: 5.7%  E*: 0.21  E90: 9.6%  E90*: 0.36  Net Benefit (prev): 0.18 | 2 |
| Bouvy et al | N=152 | Both acute and chronic HF, multicenter study Netherlands  (Distantly Related) | - Mean age 70 yrs - 35% male - LVEF not reported | All variables with p<0.10 were included in the model | - Age - Gender - DM - Renal dysfunction - Edema - Weight - SBP - Beta-blockers | 6 | Mortality at 18 months | N=51 (34%)  1011 (57%) | C-statistic = 0.80 | C-statistic: 0.65  C-statistic (Val vs. Der): -49%  Model Based C statistic: 0.83  % change in C-statistic (Val vs MBC): -53% | Calibration slope: 0.34  E: 19.9%  E*: 0.35  E90: 35.8%  E90*: 0.63  Net Benefit (prev): 0.19 | 1 |
