## Supplemental Table 2 for "Performance of Heart Failure Clinical Prediction Models: A Systematic External Validation Study"

Short title: HF prediction model external validation

Keywords: Clinical prediction model, heart failure, mortality

Acknowledgement: Research reported in this work was funded through a Patient-Centered Outcomes Research Institute® (PCORI®) Award (ME-1606-35555).

Disclaimer: The views, statements, opinions presented in this work are solely the responsibility of the author(s) and do not necessarily represent the views of  the Patient-Centered Outcomes Research Institute® (PCORI®), its Board of Governors or Methodology Committee.

Supplemental Table 2: Chronic HF CPMs with at least one validation dataset match:

| CPM Citation | Sample Size, Enrollment Years | Population | Baseline characteristics: Age, % male, mean LVEF, NYHA Class I/II | Baseline Medications | Variable Selection | Variables final model (N) | Events per variable (EPV) in Derivation Cohort | Outcome definition, N(%) in derivation cohort | Derivation c-statistic | RCT validation(s) | Number of prior validations reported in literature (through 2015) |
| --- | --- | --- | --- | --- | --- | --- | --- | --- | --- | --- | --- |
| Kraaier et al^34^ | N=861, 2002-2008 | Single-center, LVEF ≤35% receiving ICD | - 63±10 yrs - 79% male - LVEF 24% ±9% - 68% NYHA I/II | - BB: 76% - ACEi/ARB: 76% - MRA: NR - Diuretics: 74% - Statin:67% - ICD 100% - CRT 28% | Logistic regression. Backward stepwise selection with P>0.10 excluded | - Age, - LVEF - GFR, - Afib | 16 | All cause mortality at 1 year  N=63 (4.8%); | NR | HF Action, SCD HeFT, BEST, SOLVD, HEAAL | 1 |
| Bilchick et al | N=17,991, 2005-2006 | Medicare beneficiaries receiving primary prevention ICDs, Registry | - Median 73 years - 77% male - 32% LVEF ≤20% - 60% NYHA I/II | - BB: 80% - ACEi/ARB:74% - MRA: NR - Diuretic: 65% - Statin: NR - ICD: 100% - CRT: NR | Cox PH model. Final model selected based on Wald chi-square test statistic, frequency of occurrence and clinical relevance | - Age - LVEF - CKD - COPD - Diabetes - NYHA class III - AFib | 963 | All cause mortality, median f/u 4 years N=6,741 (38%)  .  For external validation, 1 year mortality used | c-statistic 0.75 | SOLVD | 1 |
| Van Rees et al | N=900, 1996-2009 | Ischemic heart disease with primary prevention ICD, single center | - 64±10 yrs; - 87% male - LVEF 29%±11% - 51% NYHA I/II | - BB: 63% - ACEi/ARB: 85% - MRA: NR - Diuretic: NR - Statin: NR - ICD: 100% - CRT: NR | Logistic regression with backward stepwise selection p<0.05 | - Age - LVEF - NYHA III-IV - Diabetes   Smoking | 23 | Death without prior ICD therapy, N=114 (13%) | c-statistic  0.73 | SCD HeFT, | 0 |
| Kramer et al | N=2717; 2001-2008 | Patients receiving ICDs at 2 centers | - 65±15 yrs - 77% male - LVEF 31%±15 - 74% NYHA I/II | NR | Logistic regression with stepwise selection p <0.1 | - Age - LVEF - Cr   Peripheral arterial disease | 31 | All cause mortality, mean follow up 3.1 years.  N=125 (14%) | c-statistic: 0.80 | HEAAL, BEST | 0 |
| Subramanian et al | N=963, 1995-1996 | Multicenter clinical trial cohort (VEST trial) | - 62±12 yrs - 78% male - LVEF 20% ±6% - 100% NYHA III/IV | NR | Logistic regression, variable selection procedure not reported | - LVEF - BUN - Lymphocytes - Cardiothoracic ratio | 43 | 1 year mortality  N=172 | c-statistic: 0.73 | SOLVD | 0 |
| Borleffs et al  (Ischemic Model) | N=704, 1996-2007 | Single-center, Patients receiving primary prevention ICD | - 64±11 yrs; - 87% male - LVEF 29%±11 - NYHA: NR | - BB: 62% - ACEi/ARB: 85% - MRA: NR - Diuretics: 72% - Statins: 82%; - ICD: 100% - CRT: 49% | Backward stepwise selection until all variables p<0.25 | - Age - LVEF - Renal function, - Diabetes - Smoking   QRS duration | NR | All cause mortality  Number of events is not reported | c-statistic: 0.81 | HF Action, HEAAL, SCD HeFT, BEST | 0 |
| Borleffs et al  (Nonischemic Model) | N=332, 1996-2007 | Single-center, Patients receiving primary prevention ICD | - 61±12 yrs - 66% male - LVEF 29%±14% - NYHA: NR | - BB: 64% - ACEi/ARB: 86% - MRA: NR - Diuretics: 82% - Statins: 32% - ICD: 100% - CRT: 70% | Backward stepwise selection until all variables p<0.25 | - Age - LVEF - Renal function   Atrial fibrillation | NR | All cause mortality  Number of events is not reported | c-statistic: 0.76 | HF Action, HEAAL, SCD HeFT, BEST | 0 |
| Goldenberg et al | N=1172, 1997-2002 | Ischemic cardiomyopathy, LVEF ≤35%, MADIT-II trial cohort | - Median age 65 yrs - 84% male - Median LVEF 25% - 76% NYHA I/II | - Diuretics: 74% - ICD: 50% | Best-subset proportional-hazards regression | - Age - BUN - NYHA Class - Atrial fibrillation - QRS duration | 36 | All cause mortality  N=183 (16%) | NR | BEST, SCD-HeFT, SOLVD | 0 |
| Alehagen et al | N=510,1996 | Swedish primary care population with symptoms possibly consistent with HF | - 73±6 yrs - 52% male - 14% LVEF <40% - 87% NYHA Class I/II | - BB: 40% - ACEi/ARB: 33% - MRA: NR - Diuretics: 45% - Statins: NR - ICD: NR - CRT: 0 | Cox PH regression. Variable selection not reported | - Age - Gender - NYHA Class - Diabetes | 18 | CV Mortality  N= 71 (14%) | c-statistic: 0.75 | BEST, DIG, HEAAL, TOPCAT | 0 |
| Bouvy et al | N=152, enrollment years not reported | Both acute and chronic HF, multicenter study Netherlands | - Mean age 70 yrs - 35% male - LVEF not reported - 45% NYHA Class I/II | - BB: 39% - ACEi/ARB: 82% - MRA: 35% - Diuretics: 100% - Statins: 25% - ICD: NR - CRT: NR | All variables with p<0.10 were included in the model | - Age - Gender - DM - Renal dysfunction - Edema - Weight - SBP - Beta-blockers | 6 | Mortality at 18 months  N=51 (34%) | c-statistic= 0.80 | BEST, SCD-HeFT, SOLVD, TOPCAT | 1 |
