## Supplemental Table 3 for "Performance of Heart Failure Clinical Prediction Models: A Systematic External Validation Study"

Short title: HF prediction model external validation

Keywords: Clinical prediction model, heart failure, mortality

Acknowledgement: Research reported in this work was funded through a Patient-Centered Outcomes Research Institute® (PCORI®) Award (ME-1606-35555).

Disclaimer: The views, statements, opinions presented in this work are solely the responsibility of the author(s) and do not necessarily represent the views of  the Patient-Centered Outcomes Research Institute® (PCORI®), its Board of Governors or Methodology Committee.

Supplemental Table 3: Chronic HF CPM CPM-Validation Results

| **CPM Citation** | **Derivation c-statistic** | **HF Action dataset validation results** | **TOPCAT dataset validation results** | **HEAAL dataset validation results** | **SCD-HeFT dataset validation results** | **SOLVD dataset validation results** | **BEST dataset validation results** | **DIG dataset validation results** |
| --- | --- | --- | --- | --- | --- | --- | --- | --- |
| Kraaier et al^34^ | NR | c-statistic: 0.61  Model Based C statistic: 0.57  Calibration slope: 0.46  E: 1.7%  E*: 0.35  E90: 3.6%  E90*: 0.75  Net Benefit (prev): 0.12  *Related | No Match | c-statistic: 0.53  Model Based C statistic: 0.59  Calibration slope: 0.27  E: 3.9%  E*: 0.64  E90: 4.8%  E90*: 0.79  Net Benefit (prev): 0.05  *Related | c-statistic: 0.57  Model Based C statistic: 0.57  Calibration slope: 0.51  E: 3.4%  E*: 0.55  E90: 4.7%  E90*: 0.76  Net Benefit (prev): 0.12  *Related | c-statistic: 0.61  Model Based C statistic: 0.53  Calibration slope: 0.63  E: 8.5%  E*: 0.68  E90: 8.3%  E90*: 0.66  Net Benefit (prev): 0.002  *Distantly Related | c-statistic: 0.59  Model Based C statistic: 0.55  Calibration slope: 0.41  E: 9.4%  E*: 0.69  E90: 9.4%  E90*: 0.69  Net Benefit (prev): 0.01  *Distantly Related | No Match |
| Bilchick et al | c-statistic 0.75 | No Match | No Match | No Match | No Match | c-statistic: 0.62  Model Based C statistic: 0.72  Calibration slope: 0.54  E: 4.0%  E*: 0.32  E90: 5.0%  E90*: 0.40  Net Benefit (prev): 0.14  *Distantly Related | No Match | No Match |
| Van Rees et al | c-statistic=0.73 | No Match | No Match | No Match | c-statistic: 0.68  Model Based C statistic: 0.71  Calibration slope: 0.72  E: 4.9%  E*: 0.30  E90: 10%  E90*: 0.60  Net Benefit (prev): 0.29  *Distantly Related | No Match | No Match | No Match |
| Kramer et al | c-statistic: 0.80 | No Match | No Match | c-statistic: 0.54  Model Based C statistic: 0.68  Calibration slope: 0.17  E: 3.2%  E*: 0.52  E90: 10.9%  E90*: 1.79  Net Benefit (prev): 0.08  *Related | No Match | No Match | c-statistic: 0.63  Model Based C statistic: 0.69  Calibration slope: 0.71  E: 10.3%  E*: 0.70  E90: 13.6%  E90*: 0.93  Net Benefit (prev): 0.09  *Distantly Related | No Match |
| Subramanian et al | c-statistic: 0.73 | No Match | No Match | No Match | No Match | c-statistic: 0.62  Model Based C statistic: 0.70  Calibration slope: 0.59  E: 2.7%  E*: 0.23  E90: 4.4%  E90*: 0.37  Net Benefit (prev): 0.18  *Distantly Related | No Match | No Match |
| Borleffs et al  Ischemic Model | c-statistic: 0.81 | c-statistic: 0.65  Model Based C statistic: 0.68  Calibration slope: 0.58  E: 2.7%  E*: 0.52  E90: 6.6%  E90*: 1.27  Net Benefit (prev): 0.26  *Distantly Related | No Match | No Match | c-statistic: 0.60  Model Based C statistic: 0.67  Calibration slope: 0.24  E: 5.9%  E*: 0.70  E90: 6.6%  E90*: 0.79  Net Benefit (prev): 0.13  *Distantly Related | No Match | c-statistic: 0.63  Model Based C statistic: 0.68  Calibration slope: 0.48  E: 7.9%  E*: 0.48  E90: 8%  E90*: 0.48  Net Benefit (prev): 0  *Distantly Related | No Match |
| Borleffs et al  Nonischemic Model | c-statistic: 0.76 | c-statistic: 0.66  Model Based C statistic: 0.67  Calibration slope: 0.55  E: 1.1%  E*: 0.25  E90: 1.6%  E90*: 0.36  Net Benefit (prev): 0.32  *Distantly Related | No Match | c-statistic: 0.63  Model Based C statistic: 0.69  Calibration slope: 0.37  E: 1.7%  E*: 0.33  E90: 2.6%  E90*: 0.50  Net Benefit (prev): 0.21  *Distantly Related | c-statistic: 0.70  Model Based C statistic: 0.69  Calibration slope: 0.72  E: 0.7%  E*: 0.18  E90: 0.7%  E90*: 0.18  Net Benefit (prev): 0.35  *Distantly Related | No Match | c-statistic: 0.59  Model Based C statistic: 0.69  Calibration slope: 0.31  E: 5.9%  E*: 0.61  E90: 6.2%  E90*: 0.65  Net Benefit (prev): 0  *Distantly Related | No Match |
| Goldenberg et al | NR | No Match | No Match | No Match | c-statistic: 0.60  Model Based C statistic: 0.56  Calibration slope: 1.56  E: 13.8%  E*: 1.11  E90: 15%  E90*: 1.21  Net Benefit (prev): 0.04  *Related | c-statistic: 0.60  Model Based C statistic: 0.65  Calibration slope: 0.52  E: 3.4%  E*: 0.16  E90: 5.8%  E90*: 0.27  Net Benefit (prev): 0.14  *Distantly Related | c-statistic: 0.60  Model Based C statistic: 0.57  Calibration slope: 1.35  E: 4.3%  E*: 0.18  E90: 5.2%  E90*: 0.22  Net Benefit (prev): 0.15  *Distantly Related | No Match |
| Alehagen et al | c-statistic: 0.75 | No Match | c-statistic: 0.61  Model Based C statistic: 0.71  Calibration slope: 0.29  E: 4.5%  E*: 0.6  E90: 7.0%  E90*: 0.93  Net Benefit (prev): 0.10  *Distantly Related | c-statistic: 0.61  Model Based C statistic: 0.68  Calibration slope: 0.43  E: 11.7%  E*: 0.40  E90: 14.6%  E90*: 0.50  Net Benefit (prev): 0.14  *Distantly Related | No Match | No Match | c-statistic: 0.56  Model Based C statistic: 0.66  Calibration slope: 0.25  E: 21.9%  E*: 0.66  E90: 23.9%  E90*: 0.72  Net Benefit (prev): 0  *Distantly Related | c-statistic: 0.62  Model Based C statistic: 0.70  Calibration slope: 0.35  E: 21.3%  E*: 0.67  E90: 28.3%  E90*: 0.89  Net Benefit (prev): 0.13  *Distantly Related |
| Bouvy et al | c-statistic= 0.80 | No Match | c-statistic: 0.63  Model Based C statistic: 0.78  Calibration slope: 0.36  E: 17.7%  E*: 2.85  E90: 42%  E90*: 6.82  Net Benefit (prev): -0.03  *Distantly Related | No Match | c-statistic: 0.69  Model Based C statistic: 0.78  Calibration slope: 0.45  E: 14.4%  E*: 1.5  E90: 53.3%  E90*: 5.44  Net Benefit (prev): -0.05  *Distantly Related | c-statistic: 0.60  Model Based C statistic: 0.78  Calibration slope: 0.26  E: 13.6%  E*: 0.81  E90: 3.0%  E90*: 1.81  Net Benefit (prev): 0.12  *Distantly Related | c-statistic: 0.62  Model Based C statistic: 0.76  Calibration slope: 0.35  E: 10.1%  E*: 0.51  E90: 19.7%  E90*: 1.0  Net Benefit (prev): 0.17  *Distantly Related | No Match |
